# The Canadian COVID-19 Experiences Project: Design and Protocol

**DOI:** 10.1101/2021.12.24.21268387

**Authors:** Peter A. Hall, Geoffrey T. Fong, Sara C. Hitchman, Anne C.K. Quah, Thomas Agar, Gang Meng, Hasan Ayaz, Bruce P. Dore, Mohammad N. Sakib, Anna Hudson, Christian Boudreau

**Author notes:** Corresponding authors: Peter A. Hall, Ph.D., School of Public Health Sciences, University of Waterloo, 200 University Avenue West, Waterloo, ON, Canada; Geoffrey T. Fong, Ph.D., Department of Psychology, University of Waterloo, 200 University Avenue West, Waterloo, ON, Canada.

## Abstract

**Introduction:** Vaccine hesitancy and inconsistent mitigation behavior performance have been significant challenges throughout the COVID-19 pandemic. In Canada, despite relatively high vaccine availability and uptake, willingness to accept booster shots and maintain mitigation behaviors in the post-acute phase of COVID-19 remain uncertain. The aim of the Canadian COVID-19 Experiences Project (CCEP) is two-fold: 1) to identify social-cognitive and neurocognitive correlates of vaccine hesitancy and mitigation behaviors, and 2) to identify optimal communication strategies to promote vaccination and mitigation behaviors into the post-acute phase of the pandemic.

**Methods and analyses:** The CCEP is comprised of two components: a conventional population survey (Study 1) and a functionally interconnected laboratory study (Study 2). Study 1 will involve 3 waves of data collection. Wave 1, completed between 28 September and 21 October, 2021, recruited 1,958 vaccine-hesitant (49.8%) and fully vaccinated (50.2%) adults using quota sampling to ensure maximum statistical power. Measures included a variety of social cognitive (e.g., beliefs, intentions) and neurocognitive (e.g., delay discounting) measures, followed by an opportunity to view and rate a set of professionally produced COVID-19 public service announcement (PSA) videos for perceived efficacy. Study 2 employs the same survey items and PSAs but coupled with lab-based eye tracking and functional brain imaging to directly quantify neural indicators of attention capture and self-reflection in a smaller community sample. In the final phase of the project, subjective impressions and neural indicators of PSA efficacy will be compared and used to inform recommendations for construction of COVID-19 PSAs into the post-acute phase of the pandemic.

**Ethics and dissemination:** The CCEP has received ethical review and approval by the University of Waterloo Office of Research Ethics. Findings will be disseminated through peer-reviewed publications and presentations at scientific meetings.

## Background

The emergence of the COVID-19 pandemic has brought to the forefront the importance of population-wide readiness for pandemic response.[1–3] For governments, this means maintaining availability of personal protective equipment, medical supplies, treatment equipment and facilities, as well as maintaining alert and communication networks to respond to the risk in a coordinated manner. Policies and procedures that facilitate intra- and international cooperation are also important. The manner in which governments of all levels communicate with population members is critical, and determining the most effective communication strategies is particularly vital with a large amount of competition from other messaging sources [4–6]

On the level of population members, pandemic response inevitably involves a number of cognitive and behavioral requirements: absorbing information about the threat, engaging in mitigation behaviors as well as taking vaccines or other medical measures when they become available. Critical determinants of vaccine uptake and mitigation behaviors include social norms, perceived risk, and beliefs about each of these and intention strength.[4–7] In a post hoc manner, reasons for taking or not taking these precautions are also critical, and the array of SARS-CoV-2 vaccines available opens the possibilities for reasons to take or not take vaccines, but also reasons for preference of one over another.

Finally, and less readily apparent, some aspects of the brain and its information processing modes can have influence on both the receipt of information communicated about personal and population level risk from an infectious agent, and behavioral responses to it. Together, the nature of population wide health communications, psychological factors influence behavioral response of population members, and neural systems coordinating information received and behavioral response are ultimately all important for pandemic response.

The Canadian COVID-19 Experiences Project was devised as a way of testing multiple levels of pandemic response, with an eye toward creating new knowledge about the most effective public-facing pandemic mitigation communications. The project involves two functionally interconnected studies, one using a conventional population survey (Study 1) and the other (Study 2) using neuroimaging and eye-tracking paradigms to augment the findings from the initial study. Importantly, a dynamic systems approach is taken, such that neuro-cognitive and social-cognitive predictors may be outcome and/or predictor of important COVID-19 mitigation phenomena.

## Study 1: Population Survey

Study 1 of the CCEP is a prospective cohort study employing a bi-annual population surveys, examining social cognitive and neurocognitive predictors of vaccination hesitancy and COVID-19 mitigation behaviors. Methods and measures are described in detail below.

## Methods

### Participants

The population survey consists of bi-annual waves with replenishment targeting 2,000 participants (18 to 54 years of age) per wave of data collection. A critical feature of the population survey is that participants are recruited using a quota system within six geographic regions (British Columbia, Alberta, Prairies (Saskatchewan and Manitoba), Ontario, Quebec, and Atlantic (New Brunswick, Nova Scotia, Prince Edward Island, and Newfoundland and Labrador), three age groups (18-24, 25-39, and 40-54) ensuring a 50/50 balance of fully vaccinated and vaccine hesitant individuals. Eligible participants were recruited from Leger Opinion, the largest proprietary nationally representative probability-based panel in Canada. Three eligible criteria were those who have yet to receive any dose of vaccine approved by Health Canada, received only one dose of a two-dose vaccine and have not yet decided to have a second dose, or have received both doses of a two-dose vaccine at Wave 1. In subsequent waves, the fully vaccinated eligibility criterion will also include the one-dose vaccine that has been approved by Health Canada.

In Wave 1 of the survey, after removing speeders from the sample, the final total sample size is 1958 (Figure 1). 50% (*N*=983) of the sample was fully vaccinated, approximately 45% (*N*=848) unvaccinated, and 5% (n=127) had a single dose without an intention to seek a second dose. The survey was administered online in English and French. Data collection for Wave 1 of Study 1 (*N*=1958) took place between 28 September and 21 October, 2021, with additional waves scheduled for every 6-month period, e.g., Wave 2 is planned for February 2022 (Figure 2).

**Figure 1.**
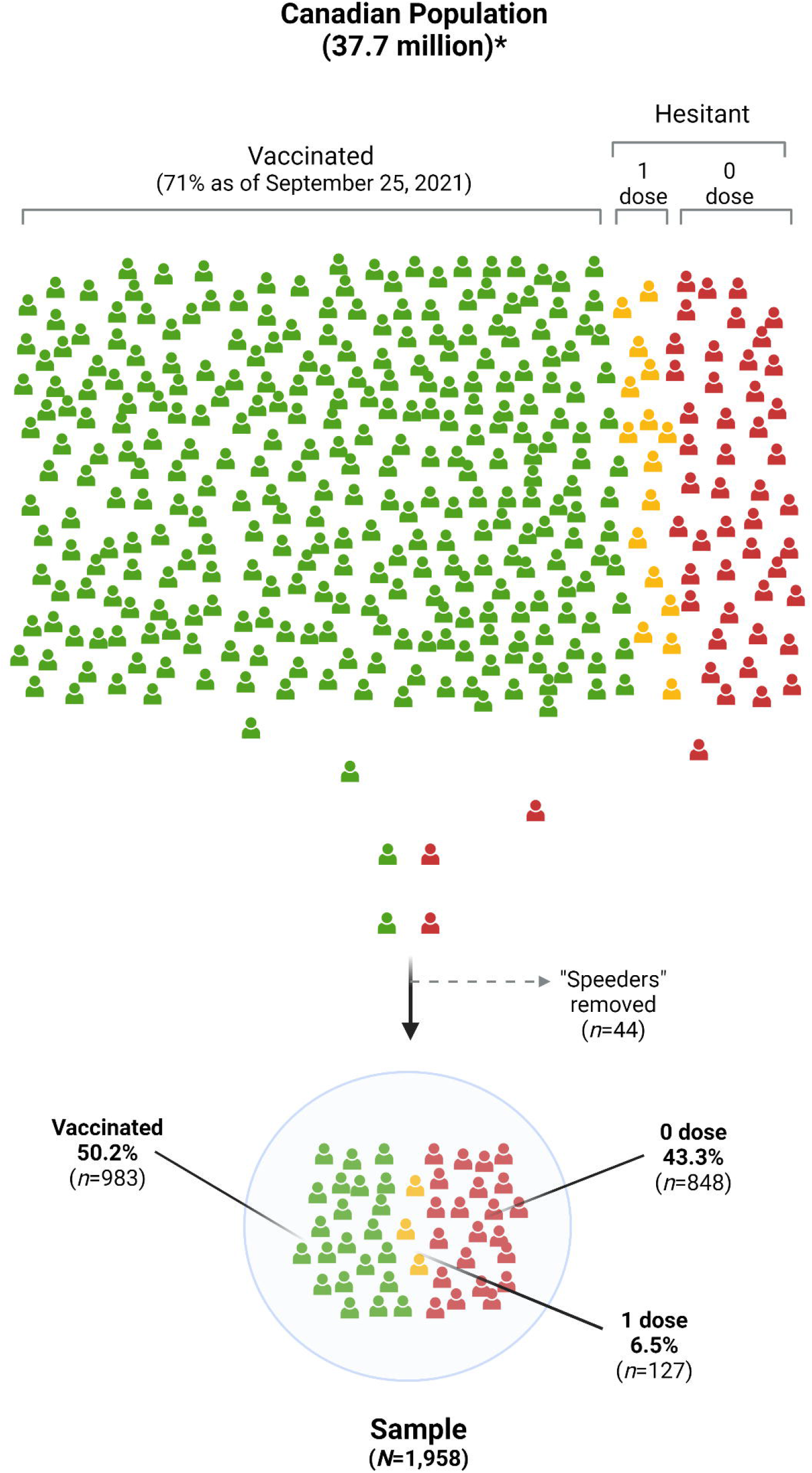
Study recruitment diagram for Wave 1 of Study 1. “1 dose”=1 vaccine dose with no intention to get a second dose (applies to 2 dose regimens only). Target recruitment for Wave 1 and future waves is set provisionally at 2,000. The sampling frame is defined as those Canadians with fixed broadband internet (estimated as 94% of the Canadian population as of 2020; Statistics Canada, 2021).

**Figure 2.**
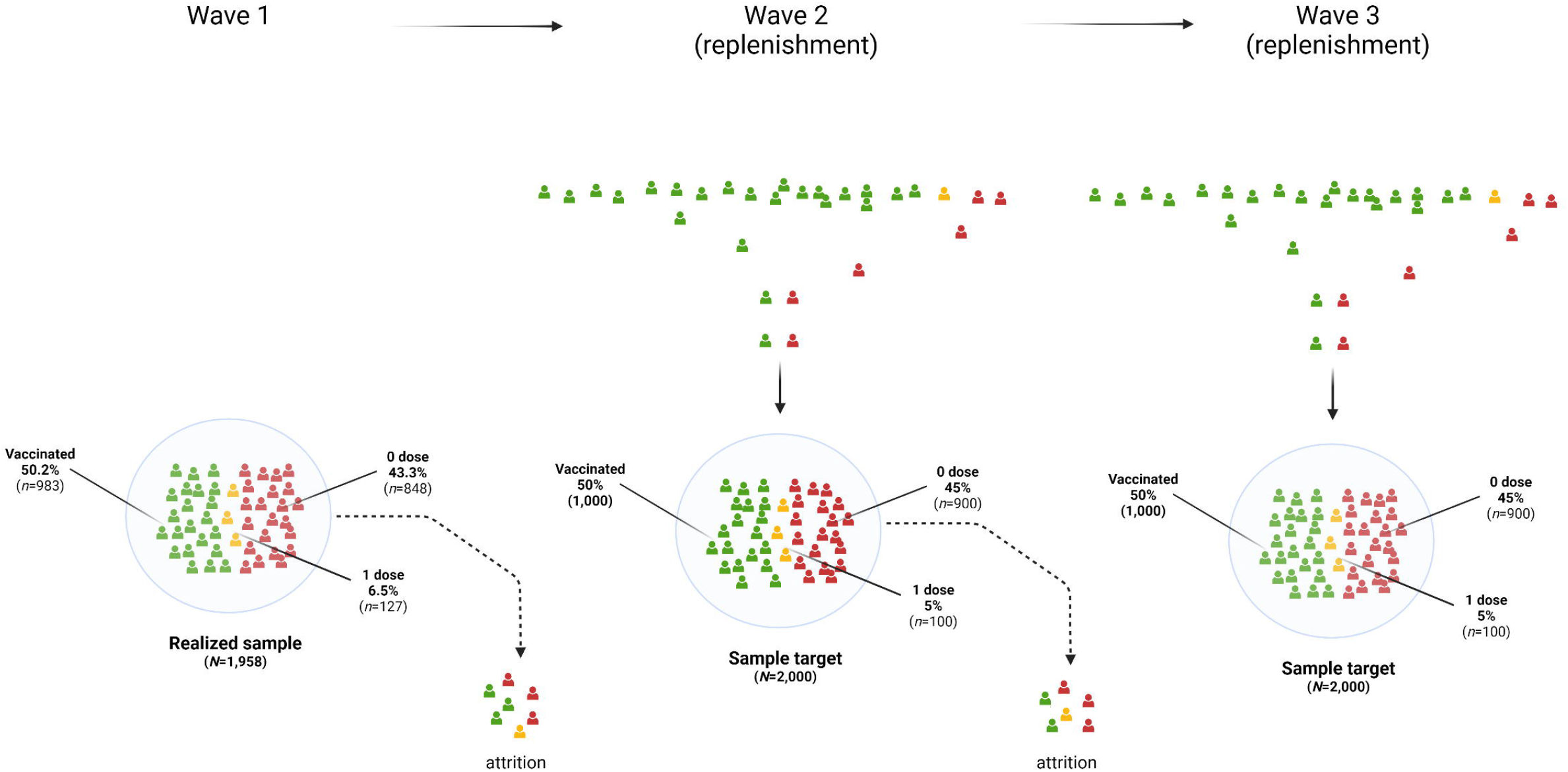
Prospective sampling and replenishment for the Canadian Covid Experiences survey from Wave 1 to Wave 3. Realized sample at Wave 1 was 2,002, reduced to 1,958 after removal of those completing the survey in times that were unrealistically rapid (“speeders”). From Wave 1 to Wave 2, and Wave 2 to Wave 3, 20% attrition is predicted, and therefore replenishment will occur at each wave to bring the sample size as close as possible to the target size of 2,000. In each wave, the target proportion of vaccine hesitant is 50%, in order to maximize statistical power.

### Sampling

A typical objective of the survey is to estimate descriptive and analytical parameters of the population. Weighting was used to represent the Canadian population based on information gathered from the sample and population figures from the Canadian census. Samples were first post stratified by geographic/language regions: Alberta, British Columbia, Manitoba + Saskatchewan, Ontario, Quebec English, Quebec French, and the Atlantic provinces (Nova Scotia, New Brunswick, Prince Edward Island, Newfoundland and Labrador). For each of the vaccinated and vaccine hesitant group separately, sampling weights were then computed using a raking procedure and calibrated to target marginal joint population distributions of the geographic/language regions, and the gender and age group combinations, based on population benchmarks in the 2016 Canadian census data and the disposition code in the sample, thus allowing generalization to the Canadian population. Two key survey statistics were computed for Wave 1 data: the response rate, defined the number of eligible respondents who completed the survey divided by the estimated number of eligible respondents that were selected/contacted (the American Association for Public Opinion Research (AAPOR) Response Rate 4 (RR4), and the cooperation rate, defined as the proportion of eligible respondents (i.e., those who have completed all eligibility questions and have been found to be eligible) who completed the survey (AAPOR Cooperation Rate 4). The cooperation rate was 94.8% and the response rate was 13.0%.

### Measures

All measures were completed online, and questions were delivered in the native language of the respondent (English or French). Question categories are described in the sections that follow:

### COVID-19 Infection History and Symptom Severity

#### SARS-CoV-2 infection history

All participants were asked, “What best describes YOUR experience with COVID-19 infection?” Those who reply “I have NOT been infected” are then asked, “How do you know that you HAVE NOT BEEN infected with COVID-19?” followed by a number of options to indicate lack of symptoms, a negative test or other reasons. Those who respond “I have been infected” to the original question are instead asked, “How do you know that you HAVE BEEN infected with COVID-19?”, followed by a number of reasons, including symptoms and a positive test result.

#### COVID-19 Disease Severity

Among those who reported being infected, respondents are then asked to describe the severity level of the symptoms using the following item stem: “How severe was your COVID-19 illness?”, with the following response options: “Not at all severe”, “Slightly severe”, “Moderately severe”, “Very severe”, “Extremely severe”. Care-level indicators of COVID-19 severity were assessed via two items: 1) hospitalization history for COVID-19, and 2) whether or not they were fitted with a ventilator during a hospitalization.

### COVID-19 Mitigation Behaviors

Participants were asked to indicate their level of consistency with respect to implementation of COVID-19 mitigation behaviours using a general measure (“How consistently do you follow the recommendations by your local or provincial public health officials about social distancing?”) as well as behavior-specific measures (“When outside your home, how consistently do you currently maintain a distance from others of at least 2 meters?”). Specific behaviors assessed include distancing (maintaining 2-meter separation), mask wearing, and hand hygiene.

### Cognitive Variables

#### Executive dysfunction

Executive function is a set of cognitive processes that constitute the ability to inhibit impulses, the ability to work with information in an “online” state, and switch flexibly back and forth between modes of responses.[8–10] Executive dysfunction was assessed using four items from the “self-restraint” subscale of the Barkley Deficits in Executive Function Scale (BDEFS).[11] Respondents were asked, “How often have you experienced each of the problems listed below during the past 6 months?” Following this, respondents were required to indicate frequency ranging from “Never or rarely” to “Very often”. Items included the following: “I am unable to inhibit my reactions or responses to events or to other people”, and “I act without thinking”, “I make impulsive comments to others”, and “I am likely to do things without considering the consequences for doing them”.

#### Delay discounting

Delay discounting is the tendency to devalue rewards disproportionately to their absolute value, as a function of delay time to receipt.[12] Those who have strong delay discounting have a tendency to impulsively reach for a lesser but immediately available reward over a much later but delayed alternative. Delay discounting was assessed using a dynamic 5-question version previously validated by Koffarnus and colleagues.[13] The module introduction started with the following instructions: “For the next 5 questions, you will be asked to choose between receiving different amounts of money at different points in time. You will see two options. You are requested to choose one of them. There are no right or wrong answers, but please take your time, answer thoughtfully, and pay careful attention to each of the options.” Participants were then given a series of monetary offers for the participant to respond to, starting with “Would you rather have $ 500 now or $ 1000 in 3 weeks?” These questions continued with the same fixed monetary amounts, but with a varied time delay for the larger amount (e.g., 1 day, 2 years). The item responses are converted to a “k” value, where higher values of k reflect higher levels of discounting of rewards as a function of time delay to receipt.

#### Time perspective

Time perspective was assessed with four survey items derived from the Time Perspective Questionnaire (TPQ)[14], two pairs of items: one pair reflecting a present orientation and a second pair reflecting a future orientation. The item pairs items may be used to assess separate values for each orientation in an orthogonal manner, or subtracted (future item pair score – present item pair score) such that higher scores on the metric reflect relatively stronger future than present orientation. Instructions for the module were as follows: “For each of the statements below, indicate your level of agreement or disagreement by using the following scale”. For each statement, participants were required to rate their level of agreement from “Strongly agree” to “Strongly disagree”. Present oriented items were as follows: “Living for the moment is more important than planning for the future.” and “I spend a lot more time thinking about today than thinking about the future.” Future oriented items were as follows: “I spend a lot of time thinking about how my present actions will have an impact on my life later on.” and “I consider the long-term consequences of an action before I do it.”

### Mood symptoms (starting in Wave 2)

#### Anxiety

Anxiety symptoms are measured using the Generalized Anxiety Disorder 7-item scale (GAD-7).[15] This scale consists of 7 items assessing a variety of symptoms of anxiety, such as: “feeling nervous, anxious, or on edge”, and “Feeling afraid, as if something awful might happen”. Responses are given on a 4-point scale, where 1 = “Not at all”, 2 = “Several days”, 3 = “More than half of the days”, 4 = “Nearly every day”. The time window for responses was 2 weeks following recovery from COVID-19.

#### Depression

Symptoms of depressions are assessed using the Centres of Epidemiological Scale – 10 item Scale (CESD-10).[16] This scale consists of 10 items assessing symptoms of low mood, lack of energy, and negative thinking styles. Sample items include “I was bothered by things that usually don’t bother me.”, and “I felt lonely.”. Responses are given on a 4-point scale, where 1 = “Rarely or none of the time”, 2 = “Some or a little of the time”, 3 = “Occasionally or a moderate amount of the time”, 4 = “All of the time”. The time window for responses was 1 week following recovery from COVID-19.

#### Agitation

Agitation is assessed using three items custom designed for the survey. Respondents are asked to indicate how often they have experienced the following symptoms during the week following recovery from COVID-19: ““I felt agitated.”, “I lashed out at other people in a way that was not like me.” “I was much more irritable than usual.” Responses are given on a 4-point scale, where 1 = “Rarely or none of the time”, 2 = “Some or a little of the time”, 3 = “Occasionally or a moderate amount of the time”, 4 = “All of the time”. The time window for responses was 1 week following recovery from COVID-19.

### Social Cognitive Variables

#### Social Norms

Social norms have been identified as being potentially important determinants of intention to perform COVID-19 mitigation behaviors and mitigation behaviors themselves.[17] Several types of norms are assessed in the survey, including descriptive, injunctive and dynamic norms.

##### Descriptive Norms

A descriptive norm is the perceived number of people that someone believes engage in a behaviour.[18] Respondents were asked about their perceived descriptive norms of vaccination, mask wearing, social distancing, and handwashing among: 1) their five most important people, 2) their province, and 3) in Canada. Respondents were first asked to list the five most important people in their life. They were then asked to report the vaccination status of their five most important people (fully vaccinated, one vaccine, no vaccines). Finally, respondents were asked to estimate what % of people in their province and what % of people in Canada are fully vaccinated or will be fully vaccinated in the next four months. For social distancing, mask wearing, and hand washing, respondents were asked to report how many of their five most important people consistently engage in the behaviour.

##### Injunctive Norms

Injunctive norms are people’s perspectives about what others believe.[19] Respondents were asked whether they “Strongly agree” to “Strongly disagree” on a 5-point scale that 1) their five most important people believe that it is important to be vaccinated, 2) people in their province believe that it is important to be vaccinated, and 3) that people in Canada believe that it is important to be vaccinated. They were then asked the same sequence of questions about social distancing, wearing masks, and washing hands.

##### Dynamic Injunctive Norms

Dynamic injunctive norms are people’s beliefs about how they think the beliefs of others are changing over time.[20] Respondents were asked which statement best describes how the beliefs about vaccines of their five most important people, people in their province, and people in Canada have changed in the last four months, with the following response options: 1) more of them believe that it is important to be fully vaccinated, 2) there is no change in the number that believe it is important to be fully vaccinated, and 3) fewer of them believe that it is important to be fully vaccinated. They were then asked questions about how the beliefs about social distancing, wearing masks, and hand hygiene of their five most important people, people in their province, and people in Canada had changed in the last four months using similar response options.

#### Reasons

Reasons for vaccination outcomes are assessed using a three-point response scale including the following options: “No, not at all a reason for me”, “Yes, this is somewhat of a reason for me”, and “Yes, this is an important reason for me.” The source list of reasons is presented in Table 1.

**Table 1.**
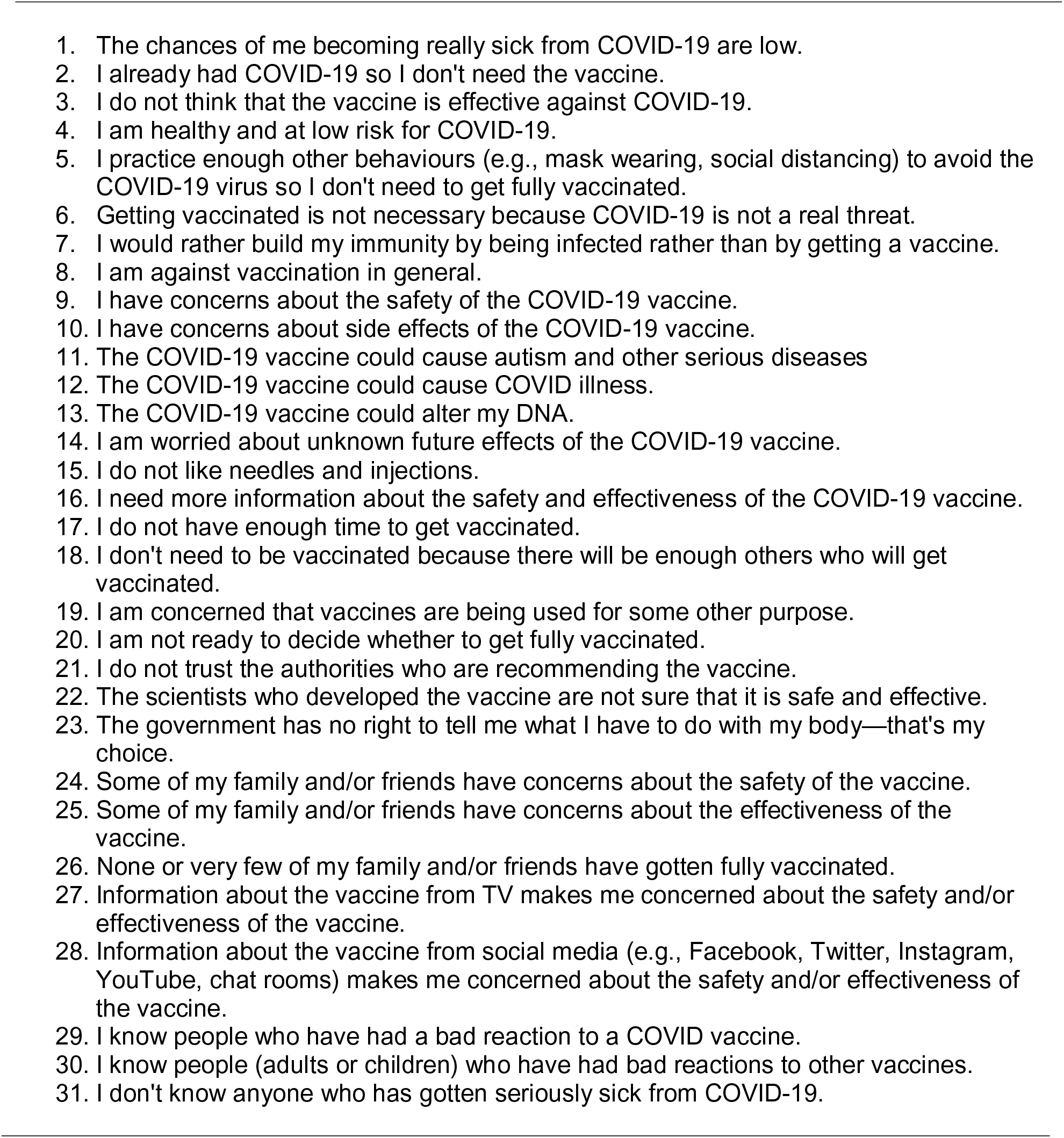
Reasons items.

##### Reasons for not getting vaccinated and not getting fully vaccinated

Respondents who were not vaccinated or had only received one dose with no plans for a second were asked reasons for not getting vaccinated or fully vaccinated.

##### Perceived reasons for not getting vaccinated

Fully vaccinated people were then presented with a list of reasons for not getting vaccinated and then asked to indicate what % of people who are not vaccinated would give those reasons.

##### Reasons for getting fully vaccinated

Among fully vaccinated people who were initially hesitant about getting vaccinated, reasons for getting fully vaccinated were asked.

#### COVID-19 Severity Beliefs and Worry

##### Severity Beliefs

Beliefs about COVID-19 severity were assessed using the following stem: “Below are a number of statements about COVID-19. For each, indicate your level of agreement.” Responses were given on a 5-point scale with response options ranging from “Strongly agree” to “Strongly disagree.” Items are presented in Table 2.

**Table 2.**
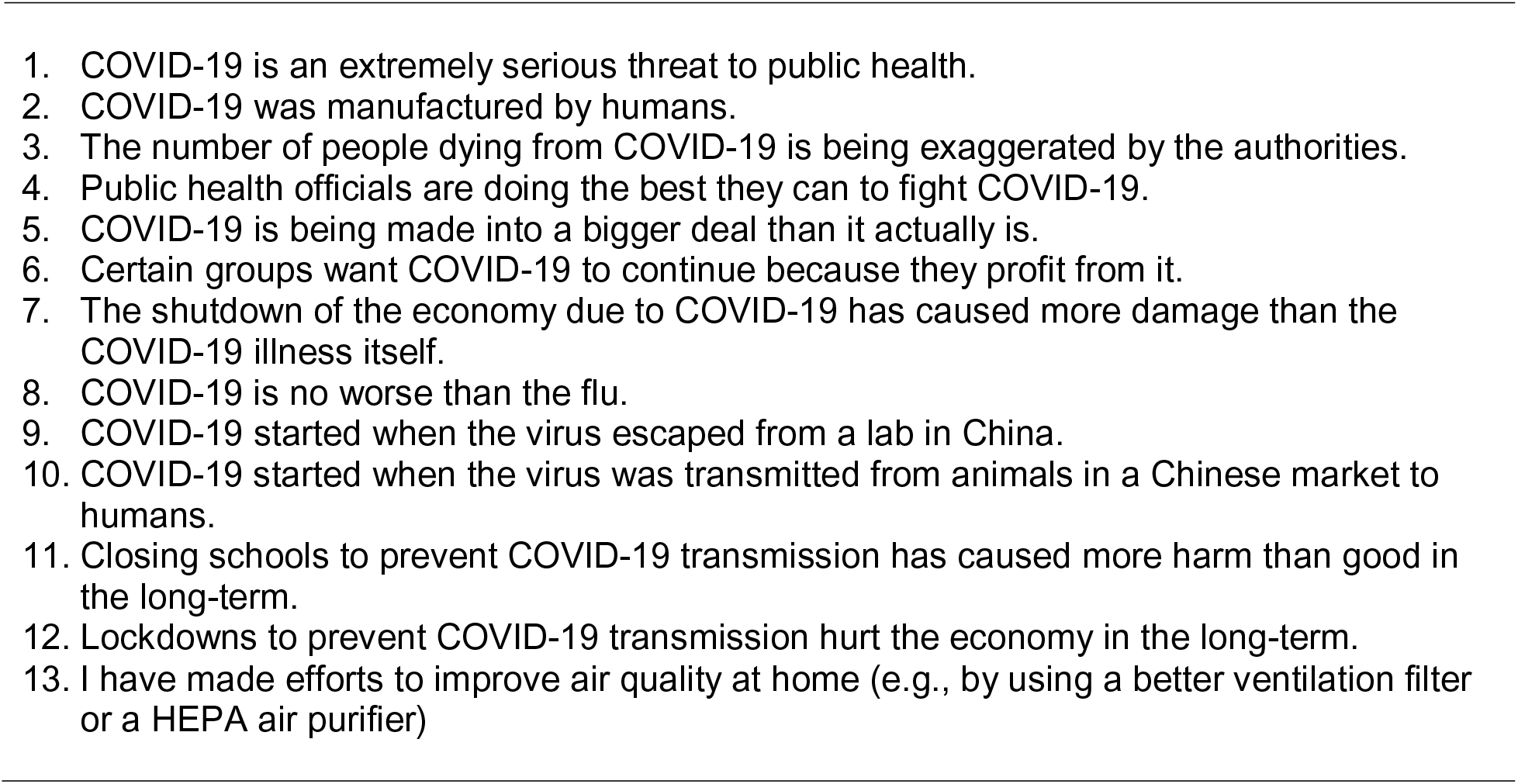
Beliefs about COVID-19 severity, origins, and mitigation measures.

##### Worry

A number of worry items were added, pertaining to the severity of COVID-19 impact on the self (e.g., “How worried are you that you will get very sick from COVID-19?”), as well as the impact on others (e.g., “How worried are you that a family member or a close friend will get very sick from COVID-19?”). Responses were given on a 5-point scale from “Not at all worried” to “Extremely worried.”

#### Perceived Effectiveness and Beliefs About Mitigation Behaviors

Participants were asked about perceived levels of effectiveness for recommended COVID-19 mitigation behaviors using the following stem: “Below are a number of statements about masks. For each, indicate your level of agreement.” Responses were given on a 5-point scale from “Strongly agree” to “Strongly disagree.” Examples of items include, “If worn properly, masks can protect the wearer from getting infected by COVID-19.”, and “Masks make it hard to breathe properly.” Behavioral targets included mask wearing, social distancing, hand hygiene, and vaccinations.

#### Vaccination Intentions

Vaccination intentions were assessed with two items, one focussed on the original vaccination and the other focussed on a hypothetical booster shot recommended in the future. The items were as follows: “What best describes your intention to get fully vaccinated in the next 4 months?” and “What best describes your intention to get an additional vaccine shot in the future (e.g., a booster shot) if the government says that it is necessary?” Response options were as follows: [I have] “No intention to get fully vaccinated”, “A very low intention”, “A low intention”, “A moderate intention”, “A strong intention”, and “A very strong intention.”

#### Political orientation, Trust in Science, Trust in Information Sources

##### Political orientation

Political orientation measures included political orientation at the provincial and national level, and where people place themselves on the political spectrum on a 7-point scale ranging from “Extremely liberal” to “Extremely conservative”, which internationally equates to “Extremely left” to “Extremely right” due to differences in the political meaning of liberal. Trust in COVID-19 information from political leaders and COVID-19 measures was also measured at the provincial and national level.

##### Trust in science

Trust in science was assessed using level of agreement for a variety of statements, listed in Table 3. Responses were on a 5-point scale from “Strongly agree” to “Strongly disagree.” Items were obtained from Nadelson and colleagues;[21] these items have been previously validated with respect to the COVID-19 context.[22]

**Table 3.**
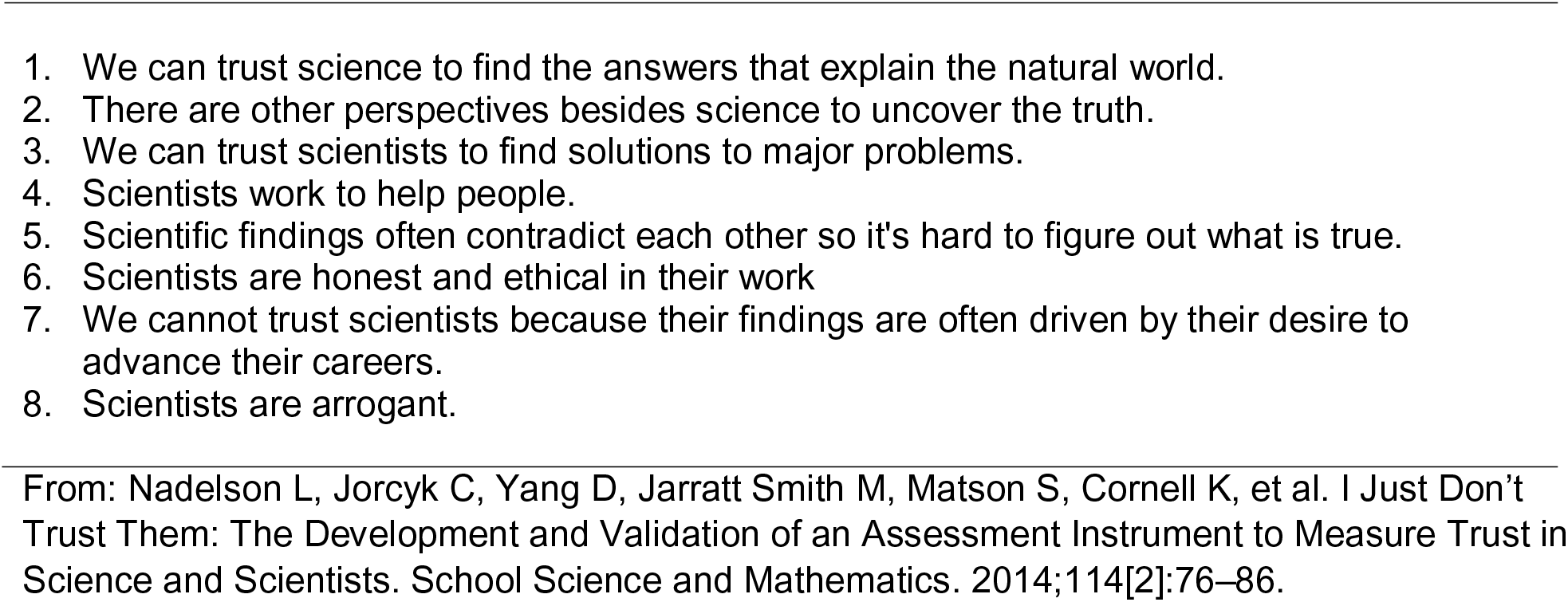
Trust in science items.

##### Information sources

Sources for obtaining COVID-19 information were assessed with the following item: “How much, if at all, do you currently get information about COVID-19 from each of the following sources?” Respondents were asked to rate on a 4-point scale from “Not at all” to “Very much” each of the following options: Church/religious group, doctor, newspapers/magazines, television, social media, or other sources.

### PSA Evaluations

A collection of public service announcements (PSAs) encouraging vaccination and COVID-19 mitigation behaviors and vaccination were professionally developed for the purpose of the CCEP. Each PSA is 25 seconds in duration, and all were produced in both official languages (i.e., English and French). Following each PSA, respondents rated the PSA on a number of evaluative dimensions, which are listed in Table 4. Most items assess the extent to which the PSAs were effective at capturing attention, motivating behavior, and encouraging reflection. Several items assessed the extent to which each PSA adhered to its intended thematic content (e.g., “How effective was this ad in getting people to think about the future benefits vs. the short-term costs of [getting vaccinated/performing mitigation behaviors]”). Rating responses were provided on a 5-point scale from “Not at all” to “Extremely.”

**Table 4.**
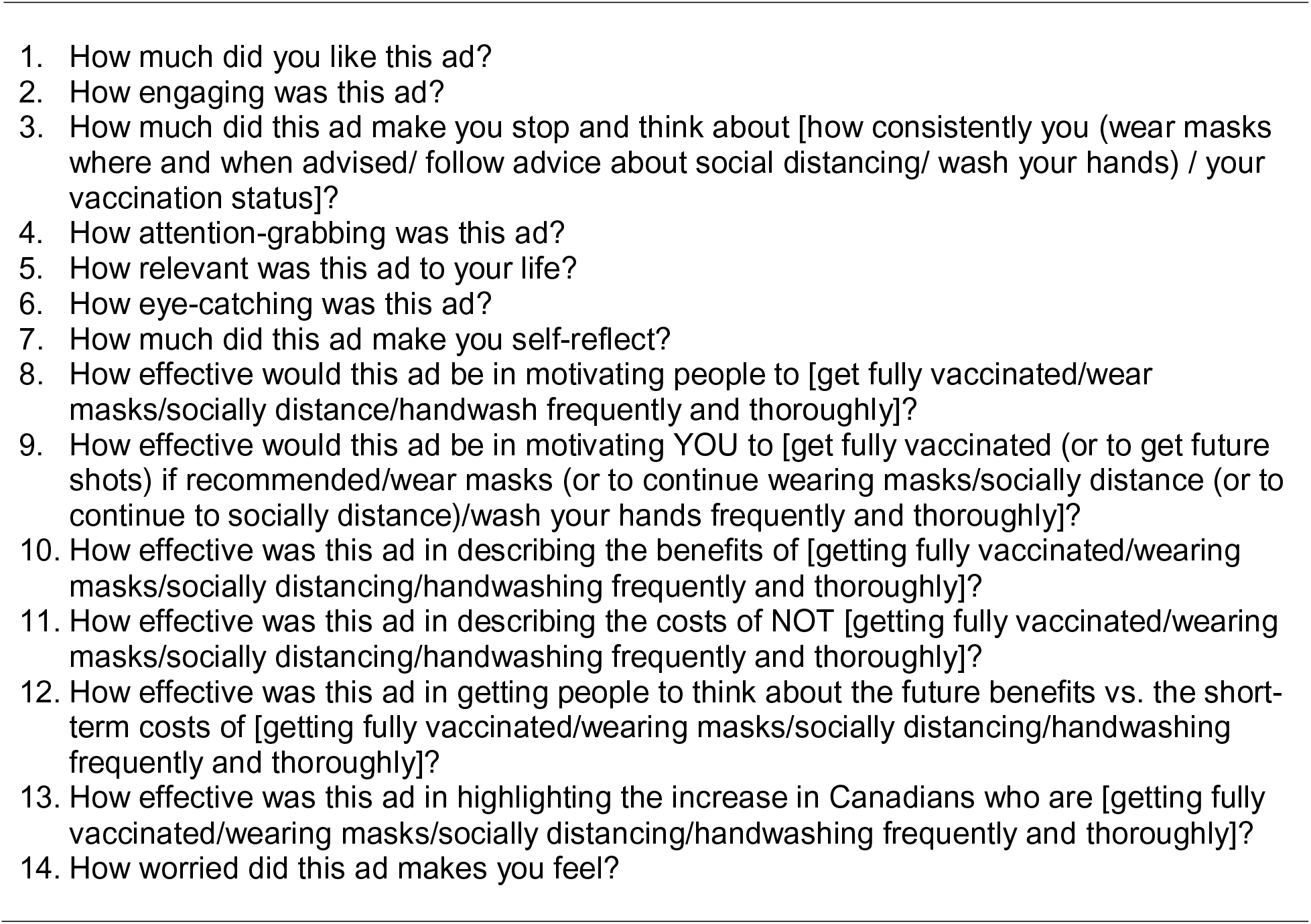
PSA Evaluation questions.

### Demographics

Age in years, gender, marital status, income, education, and ethnicity were assessed via self-report. Financial strain was assessed using two items: “In the last 30 days, because of a shortage of money, were you unable to pay any important bills on time, such as electricity, telephone or rent bills?” and “Are you currently receiving assistance or income from any federal, provincial or local programs, such as food bank use, welfare/social assistance, employment insurance, etc.?” Additional items were included to assess the extent to which individuals lost a job and/or received government benefits of various types. Demographic features of the Study 1, Wave 1 sample are presented in Table 5.

**Table 5.** Demographic features of the Study 1 Wave 1 sample.

| variable | value | Frequency | % |
| --- | --- | --- | --- |
| Gender | Male | 768 | 39.22 |
|  | Female | 1190 | 60.78 |
| Age group | 18-24 | 322 | 16.45 |
|  | 25-39 | 789 | 40.3 |
|  | 40-54 | 847 | 43.26 |
| Income | Low | 305 | 15.58 |
|  | Moderate | 436 | 22.27 |
|  | High | 1031 | 52.66 |
|  | No answer | 186 | 9.5 |
| Education | Low | 408 | 20.84 |
|  | Moderate | 710 | 36.26 |
|  | High | 810 | 41.37 |
|  | No answer | 30 | 1.53 |
| Geographic regions | AB | 245 | 12.51 |
|  | BC | 244 | 12.46 |
|  | MB+SK | 119 | 6.08 |
|  | Maritimes | 111 | 5.67 |
|  | ON | 737 | 37.64 |
|  | QC-EN | 134 | 6.84 |
|  | QC-FR | 368 | 18.79 |
| Vaccination status | I have NOT received any vaccine shot | 848 | 43.31 |
|  | Received ONE vaccine shot | 127 | 6.49 |
|  | Received TWO vaccine shots | 983 | 50.2 |
| COVID19 infection history | Not infected | 1640 | 83.76 |
|  | infected: Not at all severe | 57 | 2.91 |
|  | infected: Slightly severe | 46 | 2.35 |
|  | infected: Moderately severe | 52 | 2.66 |
|  | infected: Very severe | 18 | 0.92 |
|  | infected: Extremely severe | 4 | 0.2 |
|  | not stated | 134 | 6.84 |
|  | infected: severity not stated | 7 | 0.36 |
| Hospitalization history | Not infected | 1640 | 83.76 |
|  | not stated | 134 | 6.84 |
|  | infected not Hospitalized | 174 | 8.88 |
|  | infected hospitalized: no ventilator | 5 | 0.26 |
|  | infected hospitalized: ventilator | 5 | 0.26 |

Note: $N=1,968$ .

## Study 2: Laboratory Study

A primary outcome of Study 1 is to identify PSAs that are subjectively attention-capturing, conducive of deep self-reflection, and perceived to be effective at motiving the self and/or others. Study 2 will employ a brain-as-predictor approach to use neural signatures of attention capture and self-relevance processing to rank the same PSAs by querying the brain directly using eye tracking and brain imaging methods.[23,24] An age-stratified sample of individuals with or without a history of COVID-19 infection will view a randomized subset of PSAs from Study 1 in pseudorandomized order. During viewing, real time functional near-infrared spectroscopy (fNIRS)[25,26] will be employed to examine region-specific blood oxygenation dependent (BOLD) response as an indicator of underlying neural activity within subregions of the prefrontal cortex during PSA viewing. Increases in BOLD response in the dorsomedial prefrontal cortex will be taken as a measure of self-relevance processing.[27–29] Eye-tracking will be used to assess attention capture, via the ratio of on-versus off-target fixations during PSA viewing. PSAs will be ranked in accordance with highest self-relevance processing, highest attention capture, and their combination. We will then explore important social cognitive and neurocognitive moderators of each of these ranking orders as informed directly by the findings of Study 1. Figure 3 shows the Study 2 eye tacking and imaging protocol.

**Figure 3.**
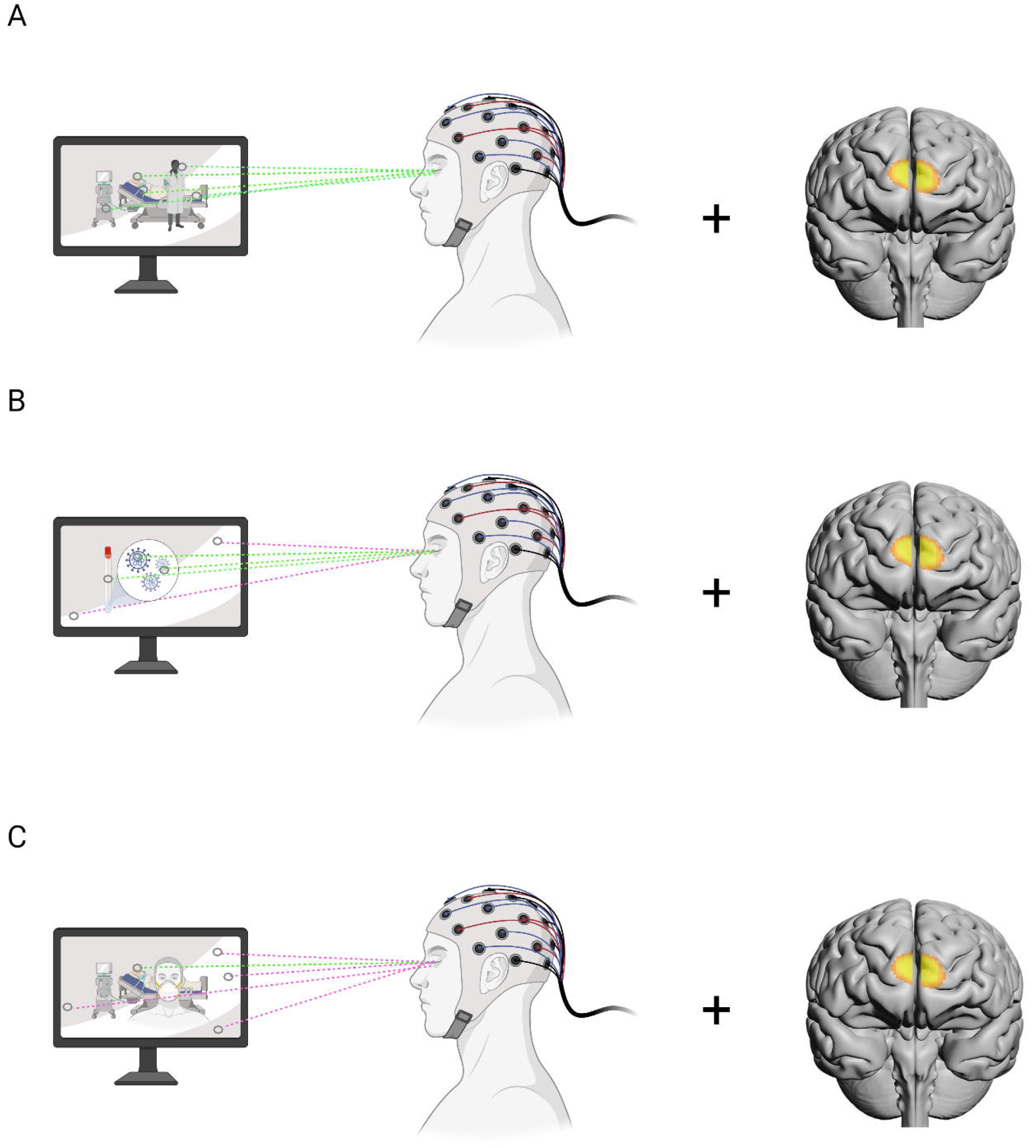
Brain imaging and eye tracking protocol for Study 2. Green dotted lines represent on-target fixations; pink dotted lines represent off-target fixations. Panel A depicts high attention capture; panel B and panel C depict moderate and low attention capture, respectively. Proportion of on-to-off target fixations will be employed as an index of attention capture for each PSA.

### Participants and Procedures

Participants will be 220 adults between the ages of 18 and 65 years, with normal or corrected-to-normal vision, and acceptable English language proficiency. Those with any diagnosed neurological disorders that might affect visual information processing or fine motor dexterity will be excluded. Following informed consent procedures, participants will complete the same online survey as per above, minus the PSA section. A PSA evaluation session will instead be completed during a laboratory visit the following day, while undergoing brain imaging using functional near-infrared spectroscopy (fNIRS) and eye-tracking. In the laboratory session, participants will be fitted with a headband containing LED light emitters and sensors, and oriented toward a screen containing a built-in eye-tracking system. Participants will view each PSA while neural response (OxyHb concentration + gaze fixations) are assessed in real time. Electronic markers will be used to identify a 5s pre-PSA baseline and two 10s epochs (baseline: second 0 to 5; early: second 6 to 15; late: second 16-25) for each PSA. Data will be analyzed such that changes from baseline to early, and baseline to late OxyHb responses will be quantified within an a-priori region of interest within the prefrontal cortex.

#### Self-relevance processing

Magnitude of self-relevance processing (deep self-reflection) will be assessed using two sets of ROIs within the prefrontal cortex (see Figure 4): dorsomedial (channels 7 and 9) and ventromedial (channels 8 and 10). These two subregions have been linked with self-relevance and evaluative processing, respectively. The dmPFC (channels 7 and 9) will be identified as the primary ROI, while the vmPFC (channels 8 and 10) are the secondary ROIs. Following data collection, each participants data will be examined visually for outlying values and anomalies. Changes in OxyHb concentrations will be the primary metric for inferring neuronal activity, with changes compared to baseline. Light intensities at 730 nm and 850 nm will be measured using COBI Studio software. Signal processing will be conducted using fnirSoft (Version 4.11). For each participant, raw light intensity will be low-pass filtered with a finite impulse response, linear phase filter with order 50 and cut-off frequency of 0.1 Hz to attenuate the high-frequency noise, respiration and cardiac cycle effects. Each channel will be checked for saturation and motion artifact contamination by means of a coefficient of variation-based assessment.[30] fNIRS data for each task block of thematically similar PSAs will be extracted using beginning and end timings based on the experiment, and hemodynamic changes for each of 16 channels during each PSA block will be calculated separately using the Modified Beer–Lambert Law with respect to the appropriate baseline. The hemodynamic response at each channel will then be averaged over time for each PSA early/late block to provide mean hemodynamic response at each channel for each block to be used in statistical analysis. PSAs thematic categories with the largest increases in OxyHb concentration from baseline (6-15s / 16-25s) in the mPFC regions (ROI, dmPFC: CH7+9 and vmPFC: CH8+10), and the largest attention effects will be identified as most compelling.

**Figure 4.**
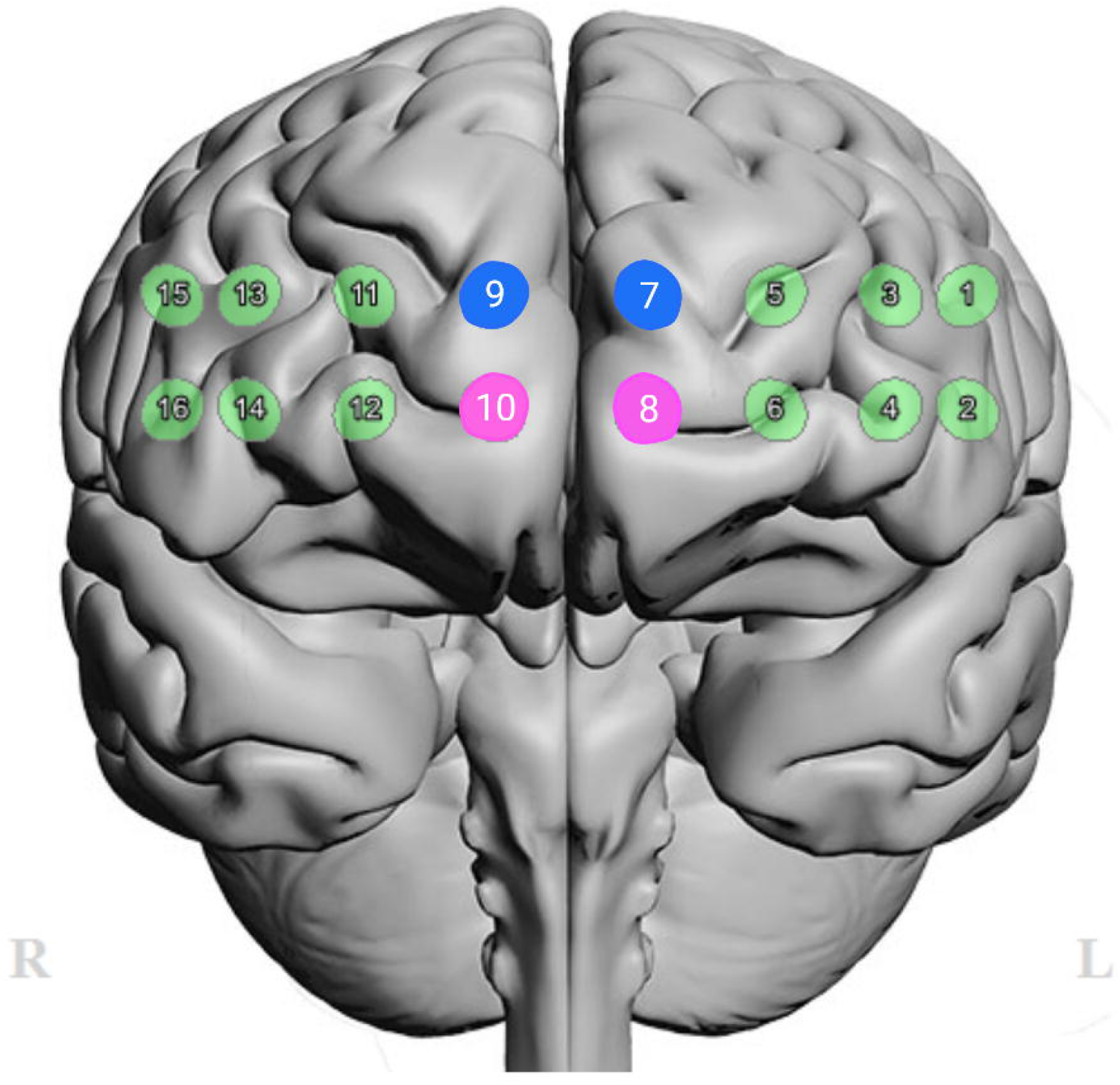
Measurement channels for functional near infrared spectroscopy (fNIRS) overlaid on an anatomical brain. Measurement channels corresponding to the dorsomedial prefrontal cortex (dmPFC) are shown in blue (channels 7 and 9). Measurement channels corresponding to the ventromedial prefrontal cortex (vmPFC) are shown in pink (channels 8 and 10). The primary metric of interest will be changes in OxyHb from baseline in each region of interest.

#### Attention capture

Attention capture will be assessed using eye tracking technology (Tobii Pro Spectrum). Eye -tracking methodology is widely used to study attentional allocation in multimedia environments.[31–33] The proportion of on target to off target fixations on a target identified within a video sample will indicate effective attention capture and engagement. PSAs developed for Study 1 will be presented as stimuli, and coupled with eye-tracking (and neuroimaging, as above) to identify degree of attentional capture on target (and neural activity indicating self-relevance processing). Participants will view each PSA from a distance of 55cm, fixed with the use of a chinrest. PSAs will be presented on a flat panel display. The eye tracking device will sample at the 300Hz frequency, and capture gaze locus throughout each PSA viewing. The target interest area (IA) will be defined separately for each PSA; and PSAs will be designed specifically to have an easily identifiable IA jittered within 20% of the centre of the frame. Fixation count percentage (proportion of total fixations in the IA) will be the primary outcome (higher% indicating more attention capture, as per past studies).

#### Statistical Analyses

dmPFC and vmPFC activations and attention capture will be used to jointly rank order PSAs, in a stepwise manner. In the first step, those PSAs with the highest proportion of fixations on the AI, will be short listed. These will then be rank-ordered with respect to neural signatures denoting self-relevance/evaluative processing, and the top 5 videos will be selected. This two-stage vetting process will be repeated for each age group. Further analyses will be undertaken to examine what factors beyond age predict attention capture (% IA fixations) and self-relevance processing (mPFC activations), and to examine links between subjective impressions and each of these dimensions. Findings gleaned from Study 1 will be compared to those from Study 2, to determine the concordance (or not) between the two information sources (i.e., subjective impressions versus attentional engagement/neural signatures). Together, these analyses will help to identify what potential mechanisms govern response to PSAs of different types, and this in turn will improve dramatically our knowledge base about how to construct captivating and thought-provoking PSAs for COVID-19 and other infectious disease threats in the future.

### Patient and Public Involvement

This investigation does not involve patients and did not involve members of the public in design or implementation

## Discussion

The CCEP is one of many studies examining predictors of vaccination and adherence in the wake of the COVID-19 crisis. For this reason, findings from this study may be considered collectively with other studies to draw the most reliable conclusions possible about the predictors of vaccine hesitancy and mitigation behavior implementation. There are, however, several features that distinguish the CCEP from other studies, including the integration of quota sampling (Study 1) and the examination of neural indicators of PSA effectiveness. The former ensures that the CCEP Study 1 survey will have maximum statistical power to identify reliable predictors of vaccine hesitancy, perhaps more so than much larger studies that do not employ quota sampling. The equal numbers of vaccinated and vaccine hesitant individuals in the sample mean that limited power will be an unlikely reason for any null predictors; conversely, we are unlikely to falsely identify significant predictors, for the same reasons. On the other hand, in Study 2, the inclusion of neurocognitive predictors and brain-related PSA evaluation are novel in the infectious disease context, if not entirely unique to our study. We will however be able to examine interactions between neurocognitive and social cognitive predictors, in part because of the wide breadth of the latter measures included. Finally, the timing of the first wave of vaccinations resulted in wide availability of vaccines in every province in Canada, ensuring that limited vaccine access is not likely to be a confound for any effects that we might observe.

Finally, a primary purpose for the CCEP was to evaluate a large pool of custom developed PSAs for promoting vaccination and COVID-19 mitigation behaviors among members of the general public. The CCEP will enable identification of effective communication approaches for use in future public-facing media campaigns for infectious disease mitigation. Combining this component with the aforementioned array of social cognitive and neurocognitive assessments, it will be possible to identify which messaging strategies work for which members of the population, as a function of social cognitive, neurocognitive and demographic subgroup membership. Among the most important objectives of the CCEP will be to identify thematic and perceptual features of PSAs that appeal differentially to those who are vaccine hesitant versus those who are already fully compliant.

Some limitations and logistical challenges remain, however. First, as the proportion of vaccinated individuals continues to rise, this subgroup will be increasingly difficult to identify and reach. In the current study context, the proportion of fully vaccinated individuals was well above 70% already at Wave 1 of data collection. However, it is anticipated that booster shots coinciding with future waves will provide an opportunity to shift focus from two vaccines to three and beyond. A further limitation is that hospitalization and other disease-related variables are self-reported. Finally, as the media becomes increasingly saturated with communications about COVID-19, and as the threat posed by the pandemic waxes and wanes, modelling temporal factors over time will become increasingly important. This may also have a disproportionate impact on Study 2, because of the sequencing of execution after the abatement of the fourth COVID-19 wave.

## Conclusions

In conclusion, the CCEP provides an opportunity to learn more about what drives vaccination and mitigation behavior during an active pandemic situation, leveraging both conventional population surveys and advanced neuroimaging techniques. What we learn in the context of COVID-19 will continue to provide important clues as to how to manage future infectious disease threats on the population level, and what factors within the social cognitive domain and neurocognitive domain might predict uptake of recommendations. Beyond this, the CCEP will provide highly novel information about what dimensions of PSA style and content are most appealing to the general public, and how this may vary systematically by vaccine hesitancy status and demographic subgroup.

## Data Availability

Not applicable (protocol)

## Informed Consent Statement

Informed consent was obtained on the web survey from all study participants prior to completing the survey.

## Ethics

The survey protocols and all materials, including the survey questionnaire, received ethical approval from the Office of Research Ethics, University of Waterloo, Canada (ORE# 43569).

## Acknowledgments

The authors would like to acknowledge and thank all those that contributed to The Canadian COVID-19 Experiences Project: all study investigators and collaborators, and the project staff at their respective institutions, namely Alkarim Bilawalla, Jessica Lee, Marie Jolicoeur-Becotte, Jamie Wheeler, Kai Jiang, Shelly Jordan, Simon Thompson, Lindsey Webster of the University of Waterloo, Andrew Mattern and Samantha Rochon of Leger Opinion.

## Author contributions

PH, GF and SH conceived the study, obtained the research funding and led the writing of the manuscript; AQ and TA participated in the design and implementation of the survey, as well as the monitoring of the data collection process; GM and CB devised and implemented the sample weighting procedures; MS and AH participated in the writing of the manuscript. HA and BD participated in Study 2 protocol design and signal processing plan.

## Patient and public involvement statement

Patients and members of the public were not involved in the design and conduct of this research.

